# Biomarkers in Hip Dysplasia: a Scoping Review

**DOI:** 10.1101/2025.10.04.25336599

**Authors:** Oscar Malmvig Sørensen, Søren Overgaard, Jens Laigaard

## Abstract

**Background:** Hip dysplasia is a congenital condition affecting around 5% of the population and is a major risk factor for secondary osteoarthritis and hip arthroplasty. While biomarkers have been widely studied in primary osteoarthritis, their role in hip dysplasia remains unclear.

Understanding biomarker profiles in this patient population may aid in the development of preventive and non-surgical treatment strategies, as well as monitoring the effects of surgical interventions and physical activity.

**Objective:** To systematically map and summarize the current evidence on biomarkers in patients with hip dysplasia.

**Methods:** This scoping review will follow the PRISMA-ScR guideline. Eligible studies include interventional and observational studies published in peer-reviewed journals reporting biomarker data in patients with hip dysplasia. Animal studies, conference abstracts, and studies not reporting hip dysplasia data separately will be excluded. We will search MEDLINE, Embase, and CENTRAL without study design or date restrictions, but limited to English-language publications. Two reviewers will independently screen studies and extract data, which will be synthesized in tables and summarized narratively.

**Dissemination:** The findings will be published in an international peer-reviewed journal and may inform research on disease-modifying treatments and prevention of osteoarthritis in hip dysplasia.

**Study registration:** This protocol is uploaded to MedRxiv.org

**Funding:** The review is investigator-initiated. Any costs in connection with the study will relate to the salary of the investigators and are provided by the department.

## Introduction

Hip dysplasia is a congenital condition with a prevalence of around 5%,^1^ and is defined as an anatomically deficient acetabulum.^2^ The condition is often present at birth and can be diagnosed and treated in early childhood.^3^ Despite systematic screening and treatment, hip dysplasia can persist or be diagnosed in adolescence or adulthood.^4^ The predominant symptom is activity-related hip pain, which leads to lower physical function and quality of life.^5^

Hip dysplasia is consistently associated with early onset osteoarthritis^4^ and need for hip arthroplasty.^6^ However, surgical treatment with hip arthroplasty carries risks such as infection, nerve injuries, and chronic postsurgical pain. Moreover, a hip arthroplasty has a limited lifespan, after which revision arthroplasty with additional risks is warranted.^7^ Thus, prevention and non-surgical treatment of osteoarthritis secondary to hip dysplasia is essential.

Biomarkers including IL-1RA, MIP-1β, RANTES, OC have in a previous study shown statistically significant differences between hip dysplasia patients and healthy controls.^4^ But investigation of biomarkers in patients with secondary osteoarthritis, including hip dysplasia have been less frequent compared to the rigorously investigated biomarkers in patients with primary osteoarthritis. It is therefore unclear, if biomarkers behave similarly in patients with primary osteoarthritis and patients with osteoarthritis following hip dysplasia. This is increasingly important with the development of disease-modifying drugs for primary osteoarthritis, but also to monitor the effect of osteotomy and physical activity. We therefore aimed to describe biomarkers in adult patients with hip dysplasia.

## Methods

This review will be reported following the PRISMA Extension for Scoping Reviews (PRISMA-ScR).^8^ This protocol will be made publicly available before initiation.

### Eligibility criteria

We will only consider studies published in peer reviewed journals. We will include all studies, both interventional and observational, that report data on biomarkers in patients with hip dysplasia. We will exclude published conference abstracts, animal studies, and studies where the data from patients with hip dysplasia is not reported separately.

### Information Sources

Sources of information are limited to the published journal articles, trial registration sites when available, attached supplementary materials, and appendices. Authors will not be contacted prior to study inclusion.

### Search Strategy

We will search the Cochrane Central Register of Controlled Trials database (CENTRAL), the Medical Literature Analysis and Retrieval System Online database (MEDLINE), and Excerpta Medica database (Embase) for eligible studies. There will be no study design or date limits on the search. We will consider only studies published in English. The search strategy will be finalized with help from an information specialist. The search strings used for all three databases will be attached in appendices of the final article.

#### Draft of PubMed search string

The result will be a search on hip dysplasia terms, plus biomarker terms, excluding studies in animals, i.e. #1 AND #2 AND #3:

##### #1 Search hip dysplasia

(hip dysplasia) OR “dysplasia of the hip” OR (DDH) OR (acetabular dysplasia) OR (Developmental dislocation of the hip) OR “developmental dysplasia of the hip” OR “secondary osteoarthritis”)

##### #2 Search biomarkers

(biomarker[MeSH Terms]) OR (joint fluid) OR (synovial fluid) OR (plasma) OR (Serum) OR (Urine) OR Collagen OR (blood) OR (Collagen Type II) OR (Collagen type I degredation neoepitope) OR (collagen type II degredation neoepitope) OR (Aggrecan*) OR (COL2) OR (C2C) OR (C1,2C) OR (Cartilage Oligomeric Matrix Protein) OR (COMP) OR (ARGS) OR (Aggrecan neoepitope fragment) OR (Matrix Metalloproteinases) OR (MMP) OR (Tissue Inhibitor of Metalloproteinases) OR (TIMPs) OR (CTX-II) OR (CTX) OR (c terminal telopeptide) OR (Keratan Sulfate) OR (Chondroitin Sulfate) OR (Hyaluronic Acid) OR (YKL-40) OR (Chitinase-3-like protein 1) OR (Fibronectin fragments) OR (ADAMTS4) OR (ADAMTS5))

##### #3 excluding animals

(all[sb] NOT (animals [MeSH Terms] NOT humans [MeSH Terms]))

### Study Records, Selection Process and Data Items

Two authors (OM & JL) will independently screen all records for eligibility in two steps: first titles and abstracts, and then full text. All records identified by either author will proceed to the next step. Disagreements regarding final inclusion are resolved by a senior author.

Study information will be charted by one author (OM), who may use automated data extraction tools (e.g., large language models such as ChatGPT (OpenAI) or Le Chat (Mistral AI)) for the first data charting draft. We will not use a pre-determined data-charting form, as we expect high variability between the included studies. Last, the charted data will be reviewed by another author (JL) to ensure validity.

### Risk of bias in individual studies

Not relevant

## Data synthesis

All included studies will be presented in summary tables. We plan to separately summarize studies assessing biomarkers in patients with hip dysplasia. The tables will comprise study information (first author, publication year) and a summary of the setting, methods, results, and conclusions. We will summarize in figures only if a substantial number of studies report the same outcome with similar estimates.

Planned list of figures and tables:

Figure 1: PRISMA flowchart^9^ of the records flow through the screening process.

Table 1: Overview of studies on biomarkers in patients with hip dysplasia

## Meta-bias

Not relevant

## Confidence in cumulative evidence

Not relevant

## Knowledge Dissemination

The resulting scoping review will be submitted for publication in an international, peer-reviewed medical journal.

## Data Availability

All data produced will be availabl eupon request to the authors, when the review is published and is.

## Notes

### Competing Interest Statement

The authors have declared no competing interest.

### Funding Statement

This study did not receive any funding

